# Epidemiology of pediatric astrovirus gastroenteritis in a Nicaraguan birth cohort

**DOI:** 10.1101/2023.08.24.23294584

**Authors:** Rebecca J Rubinstein, Yaoska Reyes, Fredman González, Lester Gutiérrez, Christian Toval-Ruíz, Kelli Hammond, Lars Bode, Jan Vinjé, Samuel Vilchez, Sylvia Becker-Dreps, Filemón Bucardo, Nadja A. Vielot

**Affiliations:** Department of Epidemiology, University of North Carolina at Chapel Hill, Chapel Hill, NC, United States; Center of Infectious Diseases, Department of Microbiology and Parasitology, Universidad Nacional Autónoma de Nicaragua—León, León, Nicaragua; Department of Family Medicine, University of North Carolina at Chapel Hill, Chapel Hill, NC, United States; Department of Pediatrics, University of California San Diego; Division of Viral Diseases, Centers for Disease Control and Prevention, Atlanta, Georgia

**Keywords:** birth cohort, astrovirus, Nicaragua, diarrhea, children

## Abstract

**Background:** Astrovirus is a leading cause of acute gastroenteritis in children worldwide. However, few prospective studies have analyzed astrovirus in community-dwelling pediatric populations in low-and-middle-income countries.

**Methods:** We assessed the incidence, risk factors, clinical characteristics, genotypes, viral coinfections and seasonality of astrovirus gastroenteritis in 443 healthy Nicaraguan children born in 2017-2018, followed for 36 months. Children were recruited from maternity hospitals and birth records in an economically-diverse neighborhood of León, the second-largest city in Nicaragua. Astrovirus-positive episodes and genotypes were identified from diarrheal specimens with reverse transcription quantitative polymerase chain reaction and Sanger sequencing.

**Results:** Of 1708 total specimens tested, eighty children (18%) experienced at least 1 astrovirus episode, and 9 experienced repeat episodes, mostly during the rainy season (May-October). The incidence of astrovirus episodes was 7.8/100 child-years (95% CI: 6.2, 9.8). Genotype-specific incidence of astrovirus also exhibited seasonality. Median age of astrovirus episode onset was 16 months (IQR 9, 23). Initial astrovirus episodes were not associated with protection against future episodes during the age span studied. Astrovirus cases were exclusively breastfed for a shorter period than uninfected children, and the human milk oligosaccharide lacto-N-fucopentaose-I was more concentrated in mothers of these children. Home toilets appeared to protect against future astrovirus episodes (HR=0.19, 95% CI 0.04-0.91). Human astrovirus-5 episodes, comprising 15% of all typed episodes, were associated with longer diarrhea and more symptomatic rotavirus co-infections.

**Conclusion:** Astrovirus was a common cause of gastroenteritis in this cohort, and future studies should clarify the role of astrovirus genotype in clinical infection severity.

## Background

Human astroviruses are an important cause of acute gastroenteritis (AGE) worldwide, detected in 2-9% of all AGE cases in children under age 3[1]. Infections are typically self-limiting with 1-4 days of watery diarrhea, fever and abdominal pain[2], and rarely, extra-gastrointestinal infection and death in immunocompromised adults and children[3,4]. Astrovirus frequently appears as a co-infection with other enteric viral pathogens, especially rotavirus, norovirus and sapovirus[1,5]. Co-infections with other pathogens have also been associated with more severe gastroenteritis symptoms and greater likelihood of experiencing diarrhea in low-and-middle-income country (LMIC) settings[1,6], where incidence, morbidity and mortality from childhood gastroenteritis is highest.

Astroviruses are non-enveloped single-stranded positive-sense RNA viruses in the family *Astroviridae*. Enteric infections are primarily caused by human astrovirus (HAstV) types 1-8 in the *Mamastrovirus* genus, MAstV-1 species[4]. Like other RNA viruses, astrovirus has a high rate of evolution and recombination[7]. Consequently, additional astroviruses, including the Melbourne (MLB) and Virginia (VA) types, were discovered with improved sequencing technology. Unlike HAstV 1-8 genotypes, the MLB and VA astroviruses are more closely related to animal astroviruses, and more commonly associated with extra-gastrointestinal infections in humans, especially neurological cases.

With few exceptions, most astrovirus studies in LMICs have been conducted in healthcare settings, limited to children with more severe clinical symptoms[1,6,9–12]. These studies could potentially over-estimate typical symptom duration in astrovirus infection. Also, children presenting to healthcare settings may be wealthier and could bias estimates of risk factors for infections, as they do not represent the population as a whole, although this bias may be less pronounced in countries with universal healthcare, including Nicaragua. Cross-sectional studies of hospitalized children are also susceptible to recall bias if caregivers misreport the timing of risk factors and clinical symptoms. Community-based cohort studies may better represent the populations at risk for astrovirus and capture the full spectrum of disease severity and risk factors. However, investigations of astrovirus in prospective birth cohorts in Central America have not been conducted in over 20 years[6,10].

In a prospective cohort study with weekly follow-up over three years, we describe the incidence and risk factors for astrovirus acute gastroenteritis (AGE) in Nicaraguan children.

## Methods

### Study sample

The Sapovirus Acute GastroEnteritis (SAGE) study is a population-based birth cohort of 444 mother-infant dyads in León, Nicaragua born between June 12, 2017 and July 31, 2018 and followed until infants were 36 months old[13]. Children were recruited from maternity hospitals and birth registries in an economically-diverse neighborhood of León, the second-largest city in Nicaragua. Within 10-14 days of birth, field staff recorded sociodemographic characteristics of the infants (e.g., sex, birth history, breastfeeding) and households (e.g., floor construction, water source, sanitation). For 36 months, children were visited in their households weekly for AGE symptoms of vomiting and diarrhea, where acute diarrhea was defined as an increase in stool frequency to 3 stools per 24-hour period or a substantial change in stool consistency, such as bloody, very loose, or watery stool. A new AGE episode was defined as symptoms preceded by three days of no symptoms[13]. At weekly visits, mothers were asked if they had breastfed the child on the previous day, and at monthly visits, mothers were asked if their child had consumed uncooked fruits or vegetables, seafood, or other foods outside the home within the previous 7 days, and whether the child had attended social gatherings or had contact with anyone experiencing diarrhea or vomiting in or outside the home within the previous 7 days[13]. All participants’ families provided informed consent for study participation and biobanking of samples for future research. All study materials were approved by the Ethical Committee for Biomedical Research at the Universidad Nacional Autónoma de Nicaragua—León (UNAN-León) (Acta #2-2017), the Institutional Review Board at the University of North Carolina at Chapel Hill (protocol #16-2079), and the Centers for Disease Control in Atlanta (project ID: 0900f3eb81c526a7)[13,14].

### Astrovirus detection and genotyping

When AGE was reported, a stool specimen was collected within two hours of defecation. Stool specimens were transported in a sterile plastic container or soiled diaper at 4°C to the Microbiology Department of UNAN-León and suspended in phosphate-buffered saline as previously described[13]. RNA was extracted from stool suspension using the The QIAamp Viral RNA Mini Kit (Qiagen, Valencia, California). Astrovirus-specific real-time PCR (RT-qPCR) reactions on the capsid gene were performed in a 96-well reaction plate using the Roche LightCycler® 96 System (Roche, Foster, CA) using primers described by Liu and colleagues. RT-qPCR was performed under the following conditions: 45°C for 10 min, 95°C for 10 min, followed by 40 cycles of 95°C for 15 s and 60°C for 30 s as described previously[15]. Samples with ct values ≤38 were considered positive for astrovirus, and the AGE episode during which the stool was collected was defined as an “astrovirus-episode”. All astrovirus-positive stool specimens were then re-amplified by conventional RT-PCR, positive samples with ct values ≤35 were Sanger-sequenced, and sequences were genotyped by comparing with astrovirus reference genotypes[16].

### Detection of viral co-infections in stool

RNA from norovirus GI, norovirus GII, sapovirus, and rotavirus were detected via RT-qPCR as described previously[15,17,18]

### Human milk oligosaccharide (HMO) identification

Manually-expelled breastmilk was analyzed for HMOs once from lactating mothers at approximately 2 months post-partum (range 1-4 months). Nineteen unique HMOs were extracted, identified, and quantified using fluorescent high-and-ultra-high-pressure liquid chromatography (HPLC-FL/UPLC) developed by the Bode Laboratory at the University of California, San Diego[19–21]. HPLC-FL/UPLC uses mass concentration to distinguish between structural isomers and quantifies ∼95% of all HMOs by concentration in breastmilk.

### Statistical analysis

To analyze the molecular epidemiology and seasonal frequency of astrovirus in our cohort, we generated a monthly epidemic curve of astrovirus episodes and an epidemic curve by genotype.

We also compared potential risk factors and exposures for astrovirus AGE among children who did and did not develop astrovirus AGE (e.g. cases and non-cases) using a Mantel-Haenszel chi-squared test for frequencies (%) and a two-sample t-test for means and standard deviations. Exclusive breastfeeding was defined as the total number of weeks in which the mother breastfed the child, and during which the child had not yet consumed other foods or liquids. Any breastfeeding was defined as the total number of weeks in which the mother breastfed the child, regardless of whether the child consumed other foods or liquids.

We determined the relative hazard (95% CI) of children experiencing an astrovirus episode by baseline risk factors using a Cox proportional hazards model. We also assessed the relative hazard (95% CI) of astrovirus AGE by time-varying risk factors and exposures using a time-varying Cox model. We also compared clinical characteristics among different astrovirus genotypes using a nonparametric Kruskal-Wallis test for continuous variables and a Fisher’s exact test of goodness-of-fit test for all categorical variables.

Next, we compared clinical and epidemiological characteristics of individuals who developed multiple astrovirus episodes versus one episode. Gastroenteritis severity was assessed using a severity score described previously[13,22] that incorporates symptoms and their duration, and receipt of intravenous fluids. Frequencies (%) of categorical variables and medians (interquartile ranges) of continuous variables were compared. We also assessed the conditional relative hazard (95% CI) of a second astrovirus episode among children who experienced an initial astrovirus episode using a Prentice-Williams-Peterson model[23]. Alpha was set at 0.05 for all inferential tests, with no adjustment for multiple comparisons.

## Results

Of 444 children enrolled in the SAGE cohort, 443 completed the study; one child dropped out within the first 5 weeks and was excluded from subsequent analyses[14]. Among 1708 specimens tested, eighty (18%) of the remaining 443 children experienced at least 1 astrovirus AGE episode, and 9 children experienced multiple episodes. The 443 children contributed 12,083 person-months of follow-up, and the incidence rate of astrovirus-specific AGE was 7.8/100 child-years (95% CI: 6.2, 9.8). The median age of astrovirus episode onset was 16 months (IQR 9, 23).

The majority of astrovirus episodes occurred during the rainy season (May-October) in the Pacific coastal region of Nicaragua[24], where León is located (Fig. 1a). No cases were documented during April, May (the start of the rainy season), or November (Fig. 1a). The largest cluster occurred between June-September, 2019, with 45 episodes detected in June alone (Fig. 1a). The median (IQR) age of episode onset among children infected in June/July 2019 was 18 months (16-20), slightly older than children infected at other time points (14 months [9–22]).

**Figure.**
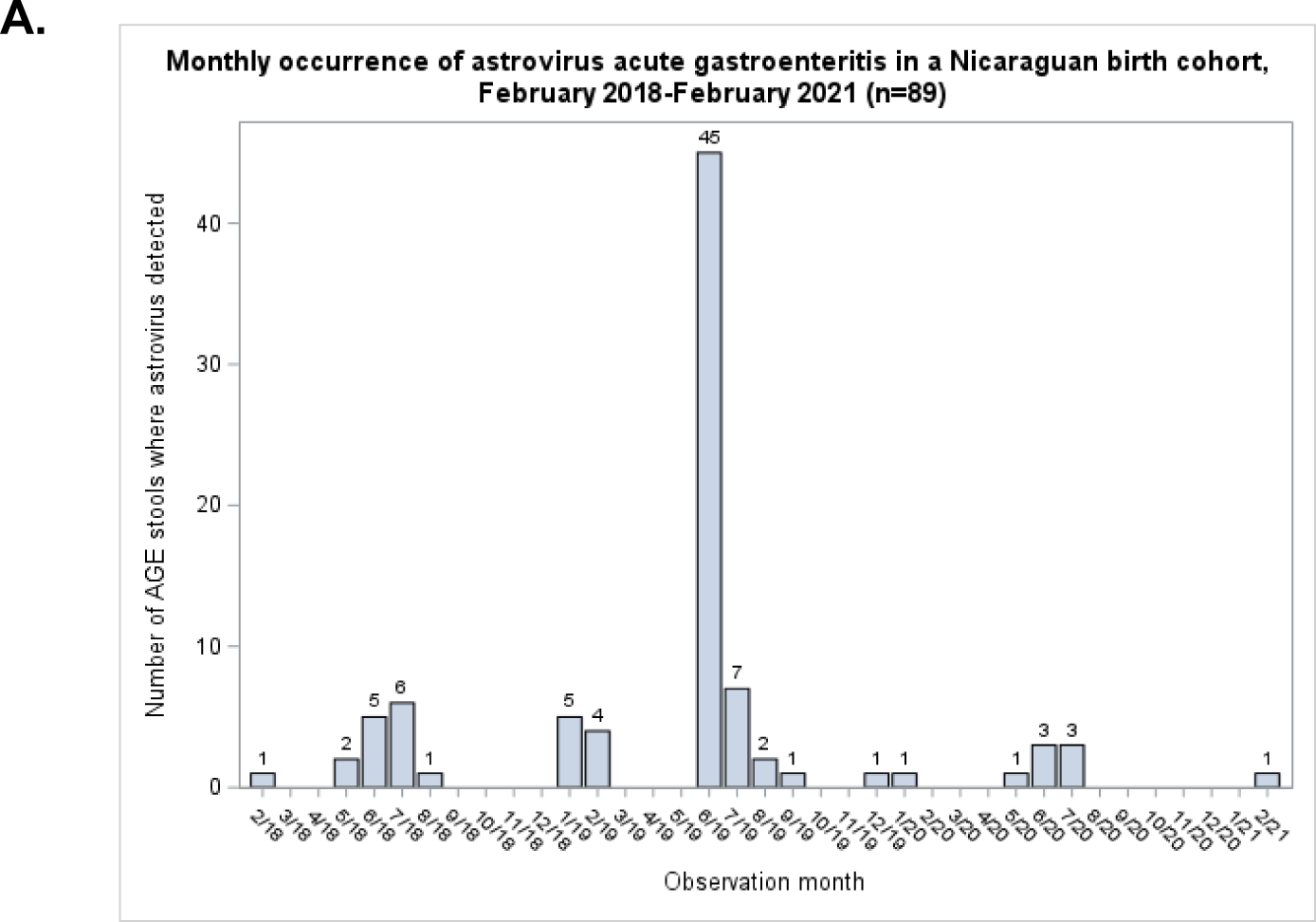
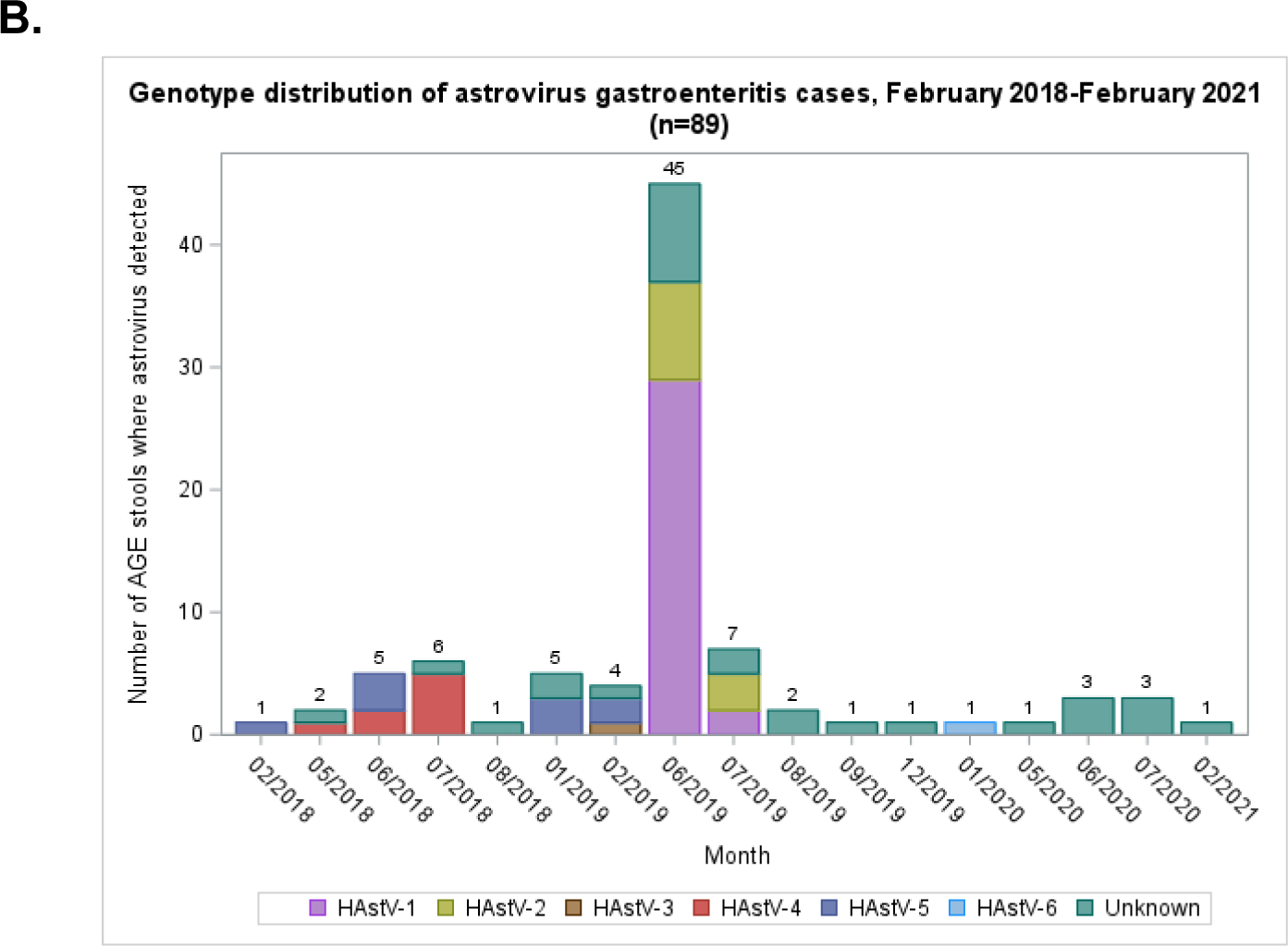
Monthly frequency of astrovirus infection a) for all astrovirus-positive gastroenteritis episodes and b) stratified by genotype in a Nicaraguan birth cohort.

The infecting genotype was determined in 61 of 89 astrovirus specimens. HAstV-1 was detected in 31 (51%) episodes (Fig 1b). Other genotypes included HAstV-2 (18%), HAstV-4 (13%), and HAstV-5 (15%). Only 1 episode of HAstV-3 and HAstV-8 was identified (1.6%).

Seasonality among the different astrovirus types was observed with HAstV-4 and HAstV-5 primarily prevalent in June-July of 2018 and January-February of 2019. In June-July, 2019HAstV-1 and HAstV-2 were primarily detected (Fig.b). Episodes of astroviruses that could not be typed were similarly distributed across all months (Fig. b).

Few characteristics differed between astrovirus cases and non-cases (Table 1). Cases were exclusively breastfed for a shorter number of weeks than non-cases, though both groups were exclusively breastfed on average for fewer than 1 study visit (Table 1). Conversely, astrovirus cases were nonexclusively breastfed for longer than non-cases (82 versus 69 weeks), though this difference was not statistically significant (Table 1). Additionally, the breastmilk consumed by astrovirus cases had a greater concentration of the α-1,2 fucosylated HMO lacto-N-fucopentaose-I (LNFP-I) (Table 1). Few characteristics were associated with time to first astrovirus episode (Table 2). Children who had contact with an individual experiencing diarrhea or vomiting in the previous week were more likely to experience an astrovirus AGE episode than those with no such contact (HR=9.08, 95% CI 5.77-14.29) (Table 2).

**Table 1.** Baseline characteristics of astrovirus AGE relative to non-cases in a birth cohort in León, Nicaragua^a^.

| Characteristic | Astrovirus AGE<br>(n=80) | Non-cases<br>(n=363) | *p*<sup>*</sup> |
| --- | --- | --- | --- |
| n (%) or mean (standard deviation) |  |  |  |
| **Birth and Genetic Characteristics** |  |  |  |
| Female sex | 33 (41%) | 184 (51%) | 0.1 |
| Mean birthweight in grams (2 missing) | 3, 148 (460.8) | 3,123<br>(438.7) | 0.7 |
| Mean age of mother years at child's birth | 23.2 (5.1) | 23.8 (5.3) | 0.3 |
| Born by vaginal delivery | 43 (54%) | 199 (55%) | 0.9 |
| **Socioeconomic Indicators & Household Sanitation** |  |  |  |
| Mother completed any secondary or higher education (12 missing) | 63 (79%) | 273 (75%) | 0.4 |
| Mother employed at time of child's birth | 13 (16%) | 60 (17%) | 0.8 |
| Presence of a toilet in the home | 62 (78%) | 261 (72%) | 0.3 |
| Presence of a dirt floor in the home | 27 (34%) | 107 (29%) | 0.5 |
| Piped municipal water in the home | 68 (85%) | 304 (84%) | 0.8 |
| **Nutrition Factors** |  |  |  |
| Mean weeks of exclusive breastfeeding | 0.07 (0.3) | 0.3 (0.4) | <0.0001 |
| Mean weeks of any breastfeeding | 82 (58.6) | 69.0 (58.2) | 0.07 |
| Human milk oligosaccharide concentrations (µg/mL) |  |  |  |
| 2_FL | 3059.9 (1644.3) | 2848.9 (1685.7) | 0.3 |
| 3FL | 508.4 (663.3) | 526.9 (475.3) | 0.8 |
| DFLac | 338.8 (346.1) | 322.2 (314.9) | 0.7 |
| 3_SL | 153.3 (115.3) | 163.4 (148.4) | 0.5 |
| 6_SL | 607.8 (294.8) | 554.5 (271.4) | 0.1 |
| LNT | 749.2 (494.6) | 761.0 (524.2) | 0.9 |
| LNnT | 171.2 (89.0) | 171.7 (95.7) | 1 |
| LNFP_I | 1354.7 (902.1) | 1114.6 (801.3) | 0.02 |
| LNFP_II | 672.1 (495.0) | 706.6 (478.3) | 0.6 |
| LNFP_III | 65.6 (63.6) | 59.2 (46.1) | 0.4 |
| LSTb | 117.5 (85.8) | 104.9 (64.6) | 0.2 |
| LSTc | 362.9 (212.8) | 320.6 (198.4) | 0.1 |
| DFLNT | 1056.5 (809.8) | 992.7 (809.2) | 0.5 |
| LNH | 172.5 (96.3) | 175.1 (111.8) | 0.9 |
| DSLNT | 228.7 (152.2) | 206.9 (127.8) | 0.2 |
| FLNH | 298.3 (151.4) | 329.1 (202.2) | 0.1 |
| DFLNH | 121.5 (123.4) | 115.5 (97.0) | 0.6 |
| FDSLNH | 250.2 (220.4) | 284.9 (232.0) | 0.2 |
| DSLNH | 282.0 (169.0) | 259.2 (148.9) | 0.2 |

\*Mantel-Haenszel chi-squared test for categorical variables; Student t-test
for continuous variables

**Table 2.**
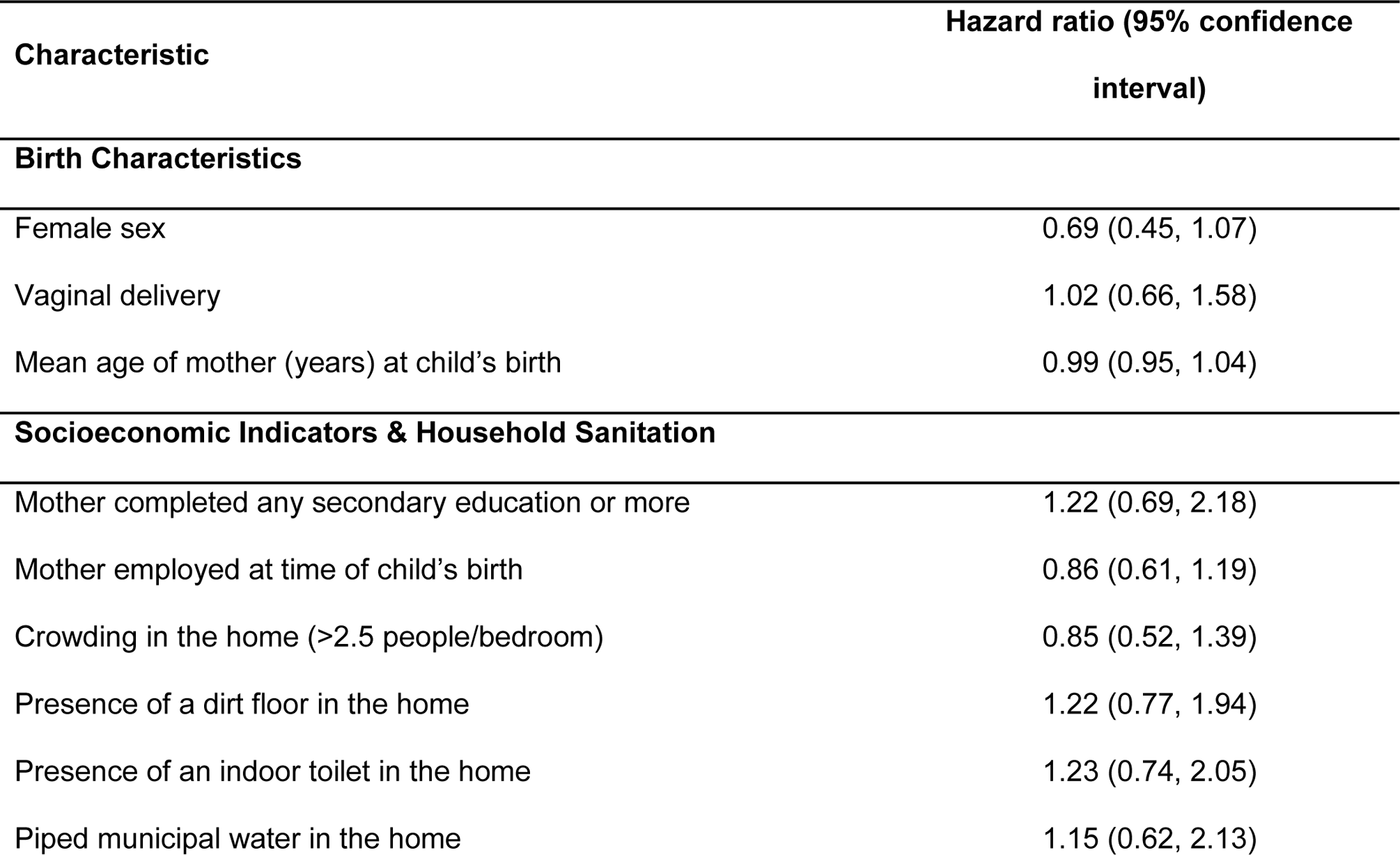
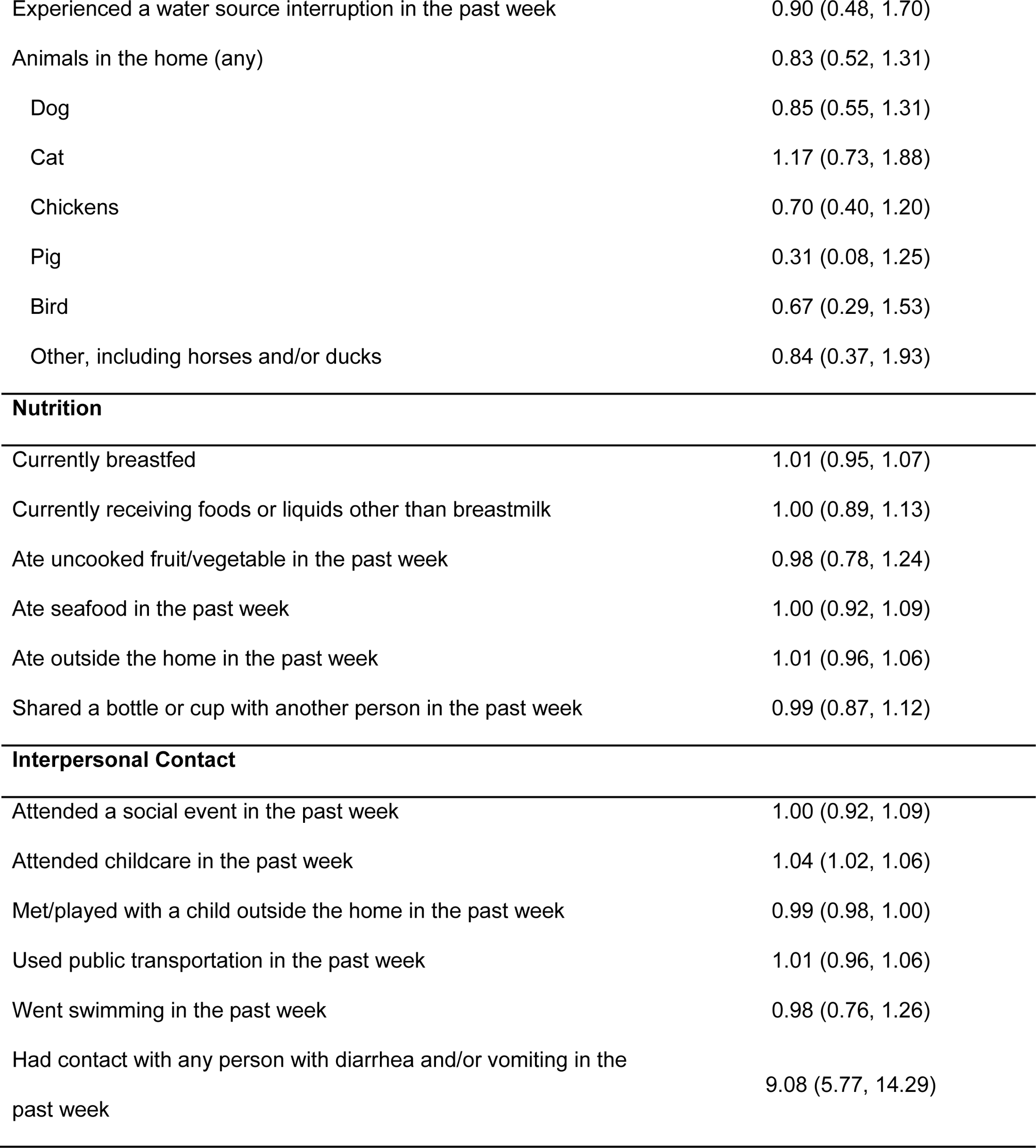
Correlates of symptomatic astrovirus-positive gastroenteritis in a Nicaraguan birth cohort (n=443)

| **Characteristic** | **Hazard ratio (95% confidence interval)** |
| --- | --- |
| **Birth Characteristics** |  |
| Female sex | 0.69 (0.45, 1.07) |
| Vaginal delivery | 1.02 (0.66, 1.58) |
| Mean age of mother (years) at child's birth | 0.99 (0.95, 1.04) |
| **Socioeconomic Indicators & Household Sanitation** |  |
| Mother completed any secondary education or more | 1.22 (0.69, 2.18) |
| Mother employed at time of child's birth | 0.86 (0.61, 1.19) |
| Crowding in the home (>2.5 people/bedroom) | 0.85 (0.52, 1.39) |
| Presence of a dirt floor in the home | 1.22 (0.77, 1.94) |
| Presence of an indoor toilet in the home | 1.23 (0.74, 2.05) |
| Piped municipal water in the home | 1.15 (0.62, 2.13) |
| Experienced a water source interruption in the past week | 0.90 (0.48, 1.70) |
| Animals in the home (any) | 0.83 (0.52, 1.31) |
| Dog | 0.85 (0.55, 1.31) |
| Cat | 1.17 (0.73, 1.88) |
| Chickens | 0.70 (0.40, 1.20) |
| Pig | 0.31 (0.08, 1.25) |
| Bird | 0.67 (0.29, 1.53) |
| Other, including horses and/or ducks | 0.84 (0.37, 1.93) |
| **Nutrition** |  |
| Currently breastfed | 1.01 (0.95, 1.07) |
| Currently receiving foods or liquids other than breastmilk | 1.00 (0.89, 1.13) |
| Ate uncooked fruit/vegetable in the past week | 0.98 (0.78, 1.24) |
| Ate seafood in the past week | 1.00 (0.92, 1.09) |
| Ate outside the home in the past week | 1.01 (0.96, 1.06) |
| Shared a bottle or cup with another person in the past week | 0.99 (0.87, 1.12) |
| **Interpersonal Contact** |  |
| Attended a social event in the past week | 1.00 (0.92, 1.09) |
| Attended childcare in the past week | 1.04 (1.02, 1.06) |
| Met/played with a child outside the home in the past week | 0.99 (0.98, 1.00) |
| Used public transportation in the past week | 1.01 (0.96, 1.06) |
| Went swimming in the past week | 0.98 (0.76, 1.26) |
| Had contact with any person with diarrhea and/or vomiting in the past week | 9.08 (5.77, 14.29) |

We also analyzed clinical characteristics and risk factors between non-astrovirus AGE episodes, initial astrovirus episodes, and subsequent astrovirus episodes (Table 3). Diarrhea was more common in initial astrovirus episodes (100%) than subsequent astrovirus episodes (67%) and non-astrovirus AGE episodes (95%). Conversely, fever was more prevalent in subsequent versus initial astrovirus episodes (49% v. 56%), while fever was less common still in non-astrovirus AGE episodes (39%) (Table 3). Nevertheless, healthcare utilization was more common among subsequent astrovirus episodes compared to initial astrovirus episodes and non-astrovirus episodes (Table 3). Additionally, children were more than twice as likely to receive zinc in a repeat astrovirus AGE episode, compared to an initial episode (67% v. 24%). Coinfection incidence was similar among initial and subsequent cases, except for sapovirus, which was more common among subsequent astrovirus episodes (22% v. 13%) (Table 3). Lastly, while having a toilet in the home appeared protective against future astrovirus AGE episodes (HR=0.19, 95% CI 0.04-0.91), no other baseline factors were associated with subsequent episodes.

**Table 3.** Clinical characteristics of astrovirus-positive gastroenteritis episodes in a birth cohort in León, Nicaragua.

|  | N (%) or Median (interquartile range) |  |  |
| --- | --- | --- | --- |
| Clinical characteristic | Non-astrovirus<br>AGE<br>(n=1367)<sup>a</sup> | First<br>episodes<br>(n=80) | Subsequent<br>episodes<sup>b</sup><br>(n=9) |
| Diarrhea | 1303 (95%) | 80 (100%) | 6 (67%) |
| Median duration in days | 6 (4-9) | 6 (3-9) | 4 (1-5) |
| Median number of stools in a 24-hour<br>period | 5 (4-7) | 5 (2) | 5 (7.5) |
| Vomiting | 352 (26%) | 24 (30%) | 3 (33%) |
| Median duration in days | 2 (2-4) | 1 (1-2) | 1 (1-3) |
| Fever | 535 (39%) | 39 (49%) | 5 (56%) |
| Median gastroenteritis severity score<br>(scale 0-15)<sup>c</sup> | 5 (4-6) | 5 (4-7) | 5 (2.5-7) |
| Received care at primary care clinic | 479 (35%) | 19 (24%) | 3 (33%) |
| Received care at emergency department | 151 (11%) | 4 (5%) | 1 (8%) |
| Admitted to hospital | 59 (4%) | 2 (3%) | 1 (8%) |
| Received zinc | 423 (31%) | 19 (24%) | 6 (67%) |
| Received intravenous fluid | 52 (4%) | 1 (1%) | 1 (8%) |
| Co-infection with other gastrointestinal<br>viruses |  |  |  |
| Norovirus GI | Not assessed (NA) | 13 (16%) | 1 (8%) |
| Norovirus GII | NA | 4 (5%) | 0 |
| Rotavirus | NA | 6 (8%) | 1 (8%) |
| Sapovirus | NA | 10 (13%) | 2 (22%) |

<sup>a</sup>Non-astrovirus AGE episodes were defined as episodes of symptomatic AGE where we tested for astrovirus RNA in stool but did not find it.
<sup>b</sup>8 children experienced a second episode, and one child experienced a third episode.
<sup>c</sup>Severity score calculated based on duration of diarrhea and or vomiting symptoms, the maximum number of stools reported per day, presence of fever, and receipt of intravenous fluid for dehydration. Adapted from the scale developed by Lee, et al.[22]

Repeat episodes from different genotypes were observed in two children. One child had HAstV-4 AGE, followed by HAstV-2 AGE over a year later, experiencing diarrhea and vomiting of 5-6 days duration in the HAstV-2 episode, but no fever. The other child was infected initially with HAstV-1, and after 14 symptom-free days, HAstV-2. The HAstV-1 episode was accompanied by diarrhea, vomiting and fever, while the HAstV-2 episode involved only diarrhea.

Most symptom prevalence, burden, and healthcare-seeking characteristics did not differ among HAstV-1, HAstV-2, HAstV-4, and HAstV-5 episodes (Table 4). We could not compare characteristics among HAstV-3 and HAstV-8, due to the small numbers of these episodes. Notably, even after eliminating extreme outliers in diarrhea duration (25 days and 88 days) among HAstV-5 episodes, diarrhea lasted significantly longer in HAstV-5 episodes (median=13.5 days, IQR 10.5, 19.5 days) than episodes with HAstV-1, HAstV-2, and HAstV-4, where median duration did not exceed 8 days (p=0.02). Additionally, the maximum diarrhea duration in HAstV-5 episodes (36 days), was more than 2 times that of all other genotypes (15 days). One-third of all children infected with HAstV-5 were also infected with wild-type rotavirus, while only 1 additional rotavirus coinfection was identified in a child with HAstV-2. This difference was statistically significant (p=0.02). The median age (IQR) at rotavirus/astrovirus co-infections was 14.6 months (12.8-16.8 months). Norovirus GI was also more common in HAstV-1 episodes (32%) and HAstV-2 episodes (18%) than in HAstV-4 or HAstV-5 episodes (0%), though this difference was not statistically significant (p=0.07). We also found that 8 (26%) of 31 HAstV-1 episodes co-occurred with sapovirus.

**Table 4.**
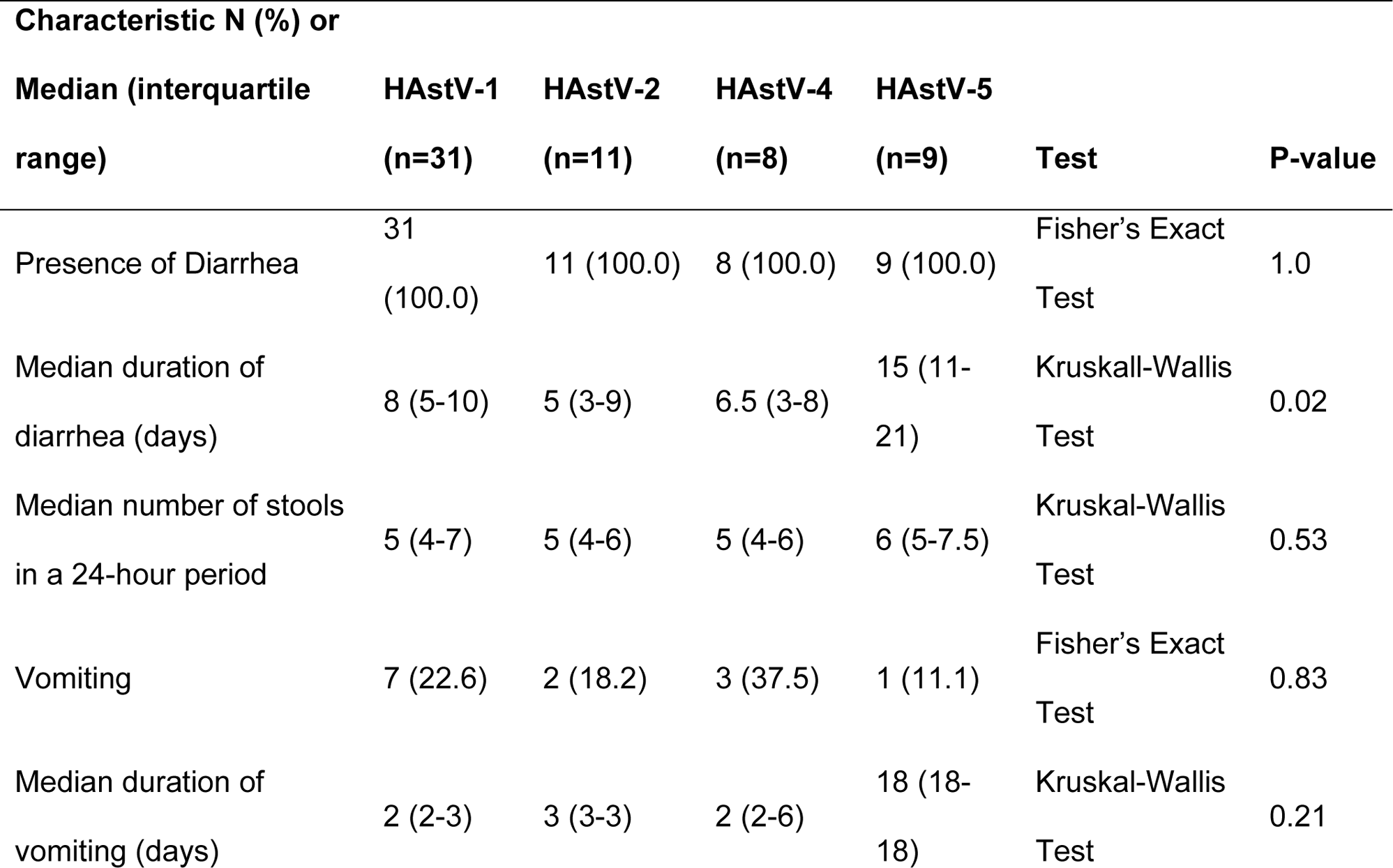
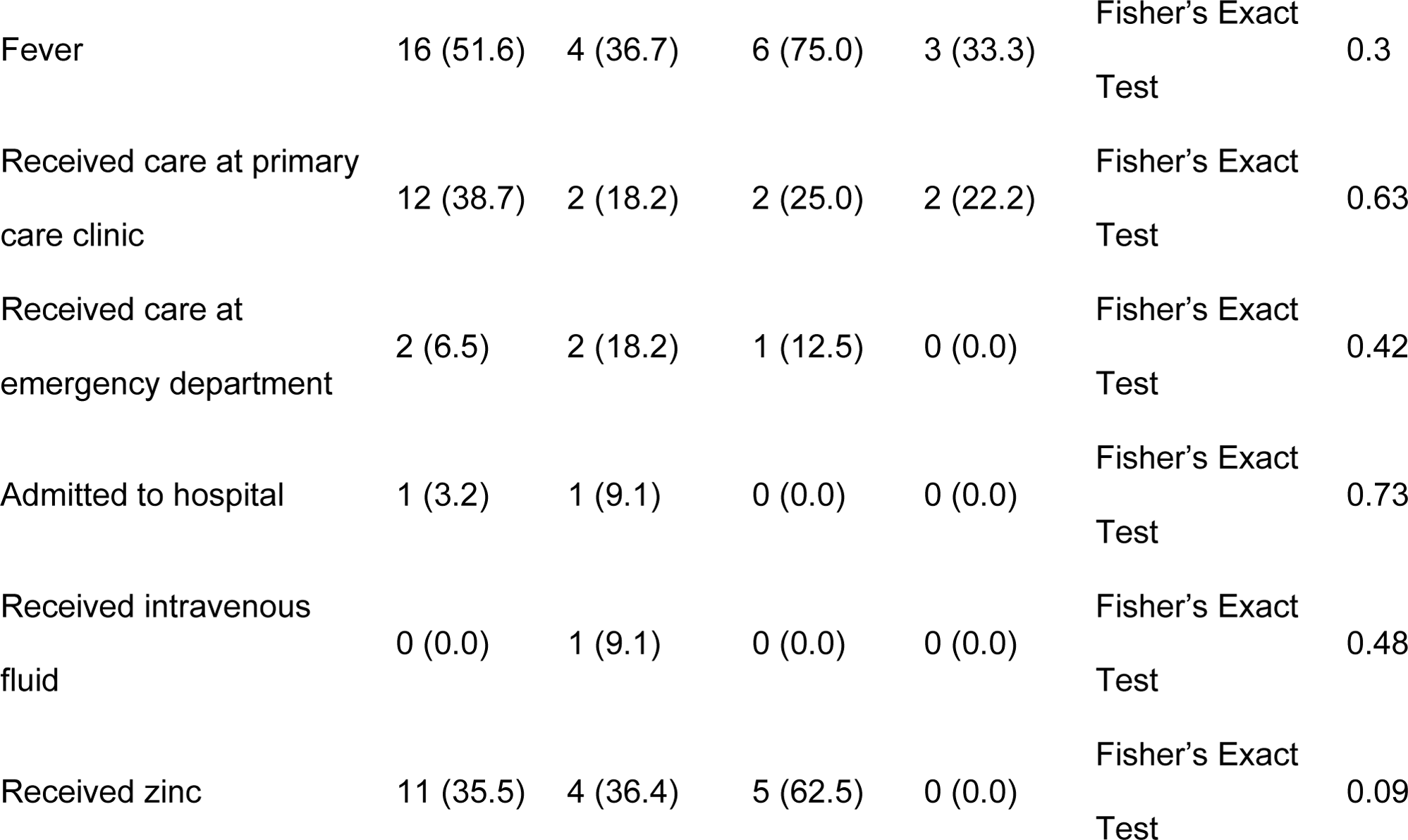
Characteristics of astrovirus-positive gastroenteritis episodes by astrovirus genotype in a Nicaraguan birth cohort (n=59).

## Discussion

Few longitudinal birth cohort studies have evaluated the risk of astrovirus in pediatric populations in low-and-middle-income (LMIC) countries[1,6,9–11,25]. Here, we present the first prospective study of symptomatic astrovirus infection burden in healthy children in Nicaragua, and only the second published study on astrovirus infection in Mesoamerica in almost 20 years[6,9,10,12,26–28]. Our findings identify potential genotype-specific differences in seasonal transmission, symptom duration, and probability of co-infections. Notably, though astrovirus has zoonotic potential, we were not able to detect non-human astrovirus genotypes with our primers, nor did we identify associations between astrovirus AGE and animals in the home. Unfortunately, we were unable to evaluate exposure to animals outside the home.

The incidence of astrovirus among AGE episodes (89/1710, 5.2%), was similar to birth cohorts from Nepal, Bangladesh, India, Pakistan, Peru, Brazil, South Africa, and Tanzania who were followed for 2 years (all 5.6%)[1,25], lower than a birth cohort from Guatemala followed for 3 years (7.3%) and a birth cohort from Mexico followed for 3 years (27.0), and higher than a birth cohort from Egypt followed for 3 years (3.5%)[1,6,11,25]. Notably, our study was conducted almost 20 years after the Egyptian, Mexican, and Guatemalan studies, during which hygiene and sanitation have improved worldwide[29]. Additionally, the molecular testing we used has higher sensitivity for pathogen detection than the ELISAs used for antigen detection in the previous studies. Nevertheless, many of our observations corroborate established evidence on astrovirus transmission. Like other tropical settings, most astrovirus episodes occurred during the rainy months of June and July[6,10,30,31], probably attributable to poorer sewage control, given astrovirus’s fecal-oral route of transmission[3]. This pattern also mirrors sapovirus and norovirus transmission in León, which tends to predominate in the rainy season[32–34].

HAstV-1 was the most common genotype in our cohort, similar to other settings[2–4,11,28]. However, HAstV-1 was not detected during all months of astrovirus transmission, appearing only in June and July 2019. Children experiencing AGE during June and July 2019 were slightly older than children infected at other time points. Although children 18 months and older may have a higher risk of astrovirus AGE than younger children[35], the small differences in age among children infected in June and July 2019 compared to other times does not likely account for the increase in infections during this time. Thus, our findings suggest that astrovirus genotypes may exhibit seasonality.

The only other identified genotype to co-circulate with HAstV-1 was HAstV-2, another commonly-detected genotype worldwide[3]. In most other months, we could only detect a single HAstV genotype, although unknown genotypes were equally distributed in almost every month of transmission. Additionally, HAstV-5 comprised almost 15% of all typed genotypes, and 11% of all initial episodes. While HAstV-5 is typically not as widespread as HAstV-1, it was common in studies from Nepal, Egypt, China, and the US, comprising 16-42% of astrovirus-positive genotypes[11,25,36,37]. Surprisingly, we also found that diarrhea lasted longer in children infected with HAstV-5. Genotype-specific astrovirus infection severity is an understudied area[38]. To our knowledge, only one other study analyzed genotype-specific symptom severity and found that HAstV-3 was associated with persistent diarrhea in previously-healthy children[39].

We also find that co-infections with wild-type rotavirus were more common among HAstV-5 episodes than among other genotypes. In Shanghai, China, patients infected with HAstV-1 and HAstV-5 were more likely to be co-infected with another enteric virus than patients infected with HAstV-2, HAstV-3, HAstV-4, or HAstV-8. However, most of these coinfections existed between norovirus and astrovirus[36], unlike our findings. While rotavirus and astrovirus coinfections commonly occur as coinfections in studies of gastroenteritis epidemiology[40,41], it is not known whether certain HAstV genotypes are more prompt to co-infections with rotavirus. Given that that astrovirus co-infections with norovirus GI (12 episodes) and sapovirus (10 episodes) were more common overall than astrovirus/rotavirus co-infections, and that norovirus and sapovirus most commonly co-occurred with HAstV-1 episodes, our data seem to suggest that different HAstV genotypes may have different propensities for co-infection with other enteric pathogens. This is another poorly-understood area that deserves further study[3].

Like Guix and colleagues[42] and Cortez and colleagues[37], we identified isolated cases of repeated astrovirus episodes of different genotypes, suggesting that different astrovirus genotypes may not offer complete cross-protective immunity[3]. Subsequent episodes in the 9 children with >1 astrovirus episode (7 with untyped episodes) were accompanied by fewer and shorter symptoms. However proportionally more children received zinc, intravenous fluids and hospital care in subsequent rather than initial episodes, conversely suggesting more clinically severe infections. Although our study was strengthened by the large number of children we followed for 3 years, we were limited by our inability to screen for asymptomatic astrovirus infections. It is possible that undetected asymptomatic astrovirus exposure early in life protected children from multiple subsequent episodes, limiting our ability to investigate whether astrovirus genotypes offer cross-protection against other genotypes. Future longitudinal studies should consider including astrovirus serology and asymptomatic astrovirus infections to clarify whether astrovirus serotypes offer cross-protection.

Astrovirus is spread through the fecal-oral route among infectious individuals[43] and through contaminated food[44], making control and prevention difficult, especially in congregate settings including schools and nursing homes[43]. Although baseline food and sanitary factors were not associated with initial astrovirus AGE in our study, we found that having a toilet in the home was protective against a subsequent astrovirus episode. Children who did not develop astrovirus were also exclusively breastfed for longer than children who became infected, consistent with findings from the 8-country MAL-ED study which identified a protective effect of prolonging exclusive breastfeeding on astrovirus infection[45]. We also found that breastmilk in mothers of infected children had greater average concentrations of the α-1,2 fucosylated HMO LNFP-I than the milk from mothers of uninfected children. A 3-country sub-analysis of the MAL-ED cohort demonstrated increased risk of astrovirus in children of mothers expressing the fucosyltransferase-3 (FUT3) gene[29], which also influences HMO composition[46]. However, LNFP-I production is driven more by expression of fucosyltransferase-2 (FUT2), which was not associated with astrovirus risk in the MAL-ED sub-cohort[29]. Although not statistically significant, we also observed that infected children were non-exclusively breastfed for longer than uninfected children. This finding is consistent with LNFP-I as a potential risk factor for astrovirus. HMOs can impact children’s health even after children are introduced to other foods[19]. The effects of HMOs on the gut microbiome could explain why LNFP-I was associated with astrovirus AGE, even though current breastfeeding was not associated with the onset of astrovirus AGE. Most of the mothers in our cohort expressed FUT2, leading to a higher concentration of LNFP-I in breastmilk (unpublished data). Children who were breastfed for longer periods would conceivably have more exposure to LNFP-I than children who were breastfed for a shorter period of time. If LNFP-I is associated with a greater incidence of astrovirus, children who were weaned later would be expected to have a higher risk of AGE. However, well-controlled observational studies and randomized trials of lab-synthesized HMOs are needed to determine whether HMOs play any role in astrovirus AGE.

Strengths of our study include its prospective design, 36-month follow-up with weekly diarrheal surveillance from birth, and its recruitment of community-dwelling children. Risk factors collected prospectively, with frequent follow-up, are less likely to be affected by recall bias. Our community-based design is potentially more generalizable and less prone to selection bias and over-estimation of astrovirus prevalence and risk factors than hospital or clinic-based studies. Additionally, we were able identify the genotype for a majority of astrovirus episodes, allowing us to compare seasonal variation in astrovirus serotype incidence and any differences in astrovirus symptom severity or co-infection incidence between serotypes. Unfortunately, we could not type all astrovirus episodes, and we did not assess for co-infections with bacteria or parasites. Thus, it is possible that symptoms observed during a so-called “astrovirus episode” were actually caused by a different pathogen. With a sample size of 443, we may have been under-powered to detect some differences, although we analyzed a larger sample than other birth cohorts in Central America[6,9]. Although we found a protective relationship between exclusive breastfeeding and astrovirus AGE, we also found that an HMO commonly found in the milk of FUT2-positive mothers was associated with an increased risk of astrovirus AGE. A future case-control study with incidence density control sampling, nested within a prospective cohort, could address these questions. Lastly, while our results are likely generalizable to urban settings on the Pacific Coast of Nicaragua, our findings may not explain astrovirus epidemiology in other areas of the country or region.

Our findings provide an estimate of astrovirus AGE risk in a community of healthy children in Central America, where few studies of astrovirus have been conducted. Our data also generate hypotheses about genotype-specific differences in astrovirus natural history and co-infections, as well as the role of HMOs in infection. Larger studies in Central America and other LMIC settings are needed to better understand the impact of astrovirus infections on child health and its prevention in regions with a high burden of diarrheal disease.

## Acknowledgments

We are very grateful to the parents and children who participated in the Sapovirus-Associated Gastro-Enterits cohort. This work would not have been possible without our dedicated field and laboratory team members: Merling Balmaceda, Vanessa Bolaños, Nancy Corea, Jhosselyng Delgado, Marvel Fuentes, Yadira Hernandez, Llurvin Madriz, Patricia Mendez, Yuvielka Martinez, Maria Mendoza, Ruth Neira, Xiomara Obando, Veronica Pravia, Yorling Picado, Aura Scott and Mileydis Soto. We also extend our thanks to the local Ministry of Health office in León and the staff of the Perla Maria Norori Health Center for their support in recruiting study participants. We also thank Mark Sotir for reviewing our manuscript.

## Data availability statement

The raw data supporting the conclusions of this article will be made available by the authors, without undue reservation.

## Author contributions

RJR, NAV, FB, SBD, JV, SV, YR, and CTV contributed to the conception and design of the study. YR, FG, LG, CTR, LB, JV, SV, FB, and NAV generated and analyzed the data. NAV, KH, and RJR performed the statistical analysis. RJR wrote the first draft of the manuscript. All authors contributed to the article and approved the submitted version. S.V, L.G, C.T-R and Y.R are former researchers from their cited primary affiliation. Credits are given because the majority of the research was performed while affiliated there.

## References

1. Olortegui MP, Rouhani S, Yori PP, et al. Astrovirus Infection and Diarrhea in 8 Countries. Pediatrics. 2018; 141(1):e20171326.

2. Bosch A, Guix S, Pintó RM. Epidemiology of Human Astroviruses. In: Schultz-Cherry S, editor. Astrovirus Res Essent Ideas Everyday Impacts Future Dir [Internet]. New York, NY: Springer; 2013 [cited 2020 Apr 1]. p. 1–18. Available from: 10.1007/978-1-4614-4735-1_1

3. Cortez V, Meliopoulos VA, Karlsson EA, Hargest V, Johnson C, Schultz-Cherry S. Astrovirus Biology and Pathogenesis. Annu Rev Virol. 2017; 4(1):327–348.

4. Vu D-L, Cordey S, Brito F, Kaiser L. Novel human astroviruses: Novel human diseases? J Clin Virol. 2016; 82:56–63.

5. Walter JE, Mitchell DK. Astrovirus infection in children. Curr Opin Infect Dis. 2003; 16(3):247–253.

6. Cruz JR, Bartlett AV, Herrmann JE, Cáceres P, Blacklow NR, Cano F. Astrovirus-associated diarrhea among Guatemalan ambulatory rural children. J Clin Microbiol. 1992; 30(5):1140–1144.

7. Babkin IV, Tikunov AY, Zhirakovskaia EV, Netesov SV, Tikunova NV. High evolutionary rate of human astrovirus. Infect Genet Evol. 2012; 12(2):435–442.

8. Roach SN, Langlois RA. Intra- and Cross-Species Transmission of Astroviruses. Viruses. 2021; 13(6):1127.

9. Walter JE, Mitchell DK, Guerrero ML, et al. Molecular Epidemiology of Human Astrovirus Diarrhea among Children from a Periurban Community of Mexico City. J Infect Dis. 2001; 183(5):681–686.

10. Maldonado Y, Cantwell M, Old M, et al. Population-Based Prevalence of Symptomatic and Asymptomatic Astrovirus Infection in Rural Mayan Infants. J Infect Dis. 1998; 178(2):334–339.

11. Naficy AB, Rao MR, Holmes JL, et al. Astrovirus Diarrhea in Egyptian Children. J Infect Dis. 2000; 182(3):685–690.

12. Diez-Valcarce M, Lopez MR, Lopez B, et al. Prevalence and genetic diversity of viral gastroenteritis viruses in children younger than 5 years of age in Guatemala, 2014–2015. J Clin Virol. 2019; 114:6–11.

13. Vielot NA, González F, Reyes Y, et al. Risk Factors and Clinical Profile of Sapovirus-associated Acute Gastroenteritis in Early Childhood: A Nicaraguan Birth Cohort Study. Ped Infect J. LWW; 2021; 40(3):220–226.

14. Reyes Y, González F, Gutierrez L, et al. Secretor status strongly influences the incidence of symptomatic norovirus infection in a genotype-dependent manner in a Nicaraguan birth cohort. J Infect Dis. 2021; :jiab316.

15. Liu J, Gratz J, Amour C, et al. A Laboratory-Developed TaqMan Array Card for Simultaneous Detection of 19 Enteropathogens. J Clin Microbiol. 2013; 51(2):472–480.

16. Noel JS, Lee TW, Kurtz JB, Glass RI, Monroe SS. Typing of human astroviruses from clinical isolates by enzyme immunoassay and nucleotide sequencing. J Clin Microbiol. 1995; 33(4):797–801.

17. Oka T, Katayama K, Hansman GS, et al. Detection of human sapovirus by real-time reverse transcription-polymerase chain reaction. J Med Virol. 2006; 78(10):1347–1353.

18. Freeman MM, Kerin T, Hull J, McCaustland K, Gentsch J. Enhancement of detection and quantification of rotavirus in stool using a modified real-time RT-PCR assay. J Med Virol. 2008; 80(8):1489–1496.

19. Lagström H, Rautava S, Ollila H, et al. Associations between human milk oligosaccharides and growth in infancy and early childhood. Am J Clin Nutr. 2020; 111(4):769–778.

20. Alderete TL, Autran C, Brekke BE, et al. Associations between human milk oligosaccharides and infant body composition in the first 6 mo of life. Am J Clin Nutr. 2015; 102(6):1381–1388.

21. Larsson MW, Lind MV, Laursen RP, et al. Human Milk Oligosaccharide Composition Is Associated With Excessive Weight Gain During Exclusive Breastfeeding-An Explorative Study. Front Pediatr. 2019; 7:297.

22. Lee G, Peñataro Yori P, Paredes Olortegui M, et al. An instrument for the assessment of diarrhoeal severity based on a longitudinal community-based study. BMJ Open. 2014; 4(6):e004816.

23. Prentice RL, Williams BJ, Peterson AV. On the regression analysis of multivariate failure time data. Biometrika. 1981; 68(2):373–379.

24. World Bank Group. Nicaragua - Climatology | Climate Change Knowledge Portal [Internet]. [cited 2023 May 4]. Available from: https://climateknowledgeportal.worldbank.org/country/nicaragua/climate-data-historical

25. Shrestha SK, Shrestha J, Andreassen AK, Strand TA, Dudman S, Dembinski JL. Genetic Diversity of Astrovirus in Children From a Birth Cohort in Nepal. Front Microbiol. 2021; 11:588707.

26. Lorenzana I, Reyes M, Ehrnst A, Hedlund K-O. Respiratory Infection and Iatrogenic Diarrhea in Honduras and El Salvador during the 1991–1992 Season. Am J Trop Med Hyg. 1996; 54(3):260–264.

27. Ortiz J, Boza AF. Molecular characterization of enteric viruses causing childhood diarrhea in Tegucigalpa, Honduras. Int J Infect Dis. 2012; 16:e198.

28. Méndez-Toss M, Griffin DD, Calva J, et al. Prevalence and genetic diversity of human astroviruses in Mexican children with symptomatic and asymptomatic infections. J Clin Microbiol. 2004; 42(1):151–157.

29. Colston JM, Francois R, Pisanic N, et al. Effects of Child and Maternal Histo-Blood Group Antigen Status on Symptomatic and Asymptomatic Enteric Infections in Early Childhood. J Infect Dis. Oxford Academic; 2019; 220(1):151–162.

30. Gamsonré Z, Bisseye C, Nitiema LW, et al. Astrovirus Gastroenteritis in Children Younger than 5 Years in Ouagadougou (Burkina Faso): Prevalence and Risks Factors Influencing Severity. Int J Trop Dis Health. 2019; :1–10.

31. Nguyen TA, Hoang L, Pham LD, et al. Identification of human astrovirus infections among children with acute gastroenteritis in the Southern Part of Vietnam during 2005–2006. J Med Virol. 2008; 80(2):298–305.

32. Vielot NA, François R, Huseynova E, et al. Association between breastfeeding, host genetic factors, and calicivirus gastroenteritis in a Nicaraguan birth cohort. Tohma K, editor. PLOS ONE. 2022; 17(10):e0267689.

33. Bucardo F, Nordgren J, Carlsson B, et al. Pediatric norovirus diarrhea in Nicaragua. J Clin Microbiol. 2008; 46(8):2573–2580.

34. Bucardo F, Reyes Y, Becker-Dreps S, et al. Pediatric norovirus GII.4 infections in Nicaragua, 1999-2015. Infect Genet Evol J Mol Epidemiol Evol Genet Infect Dis. 2017; 55:305–312.

35. Nguekeng Tsague B, Mikounou Louya V, Ntoumi F, et al. Occurrence of human astrovirus associated with gastroenteritis among Congolese children in Brazzaville, Republic of Congo. Int J Infect Dis. 2020; 95:142–147.

36. Wu L, Teng Z, Lin Q, et al. Epidemiology and Genetic Characterization of Classical Human Astrovirus Infection in Shanghai, 2015–2016. Front Microbiol. 2020; 11:570541.

37. Cortez V, Freiden P, Gu Z, Adderson E, Hayden R, Schultz-Cherry S. Persistent Infections with Diverse Co-Circulating Astroviruses in Pediatric Oncology Patients, Memphis, Tennessee, USA. Emerg Infect Dis. 2017; 23(2):288–290.

38. Padron Regalado E. Molecular Epidemiology of Viral Gastroenteritis in Hajj pilgrimage [Internet]. [Thuwal, Kingdom of Saudi Arabia]: King Abdullah University of Science and Technology; 2014. Available from: https://repository.kaust.edu.sa/handle/10754/316701

39. Caballero S, Guix S, El-Senousy WM, Calicó I, Pintó RM, Bosch A. Persistent gastroenteritis in children infected with astrovirus: Association with serotype-3 strains: Persistent Astrovirus Gastroenteritis. J Med Virol. 2003; 71(2):245–250.

40. Kumthip K, Khamrin P, Ushijima H, Maneekarn N. Molecular epidemiology of classic, MLB and VA astroviruses isolated from <5 year-old children with gastroenteritis in Thailand, 2011-2016. Infect Genet Evol J Mol Epidemiol Evol Genet Infect Dis. 2018; 65:373–379.

41. Levidiotou S, Gartzonika C, Papaventsis D, et al. Viral agents of acute gastroenteritis in hospitalized children in Greece. Clin Microbiol Infect. 2009; 15(6):596–598.

42. Guix S, Caballero S, Villena C, et al. Molecular Epidemiology of Astrovirus Infection in Barcelona, Spain. J Clin Microbiol. 2002; 40(1):133–139.

43. Thongprachum A, Khamrin P, Maneekarn N, Hayakawa S, Ushijima H. Epidemiology of gastroenteritis viruses in Japan: Prevalence, seasonality, and outbreak. J Med Virol. 2016; 88(4):551–570.

44. Iritani N, Kaida A, Abe N, et al. Detection and genetic characterization of human enteric viruses in oyster-associated gastroenteritis outbreaks between 2001 and 2012 in Osaka City, Japan. J Med Virol. 2014; 86(12):2019–2025.

45. McCormick BJ, Richard SA, Murray-Kolb LE, et al. Full breastfeeding protection against common enteric bacteria and viruses: results from the MAL-ED cohort study. Am J Clin Nutr. 2022; 115(3):759–769.

46. Bode L, Jantscher-Krenn E. Structure-function relationships of human milk oligosaccharides. Adv Nutr Bethesda Md. 2012; 3(3):383S–91S.

